# Clinical Significance of Circulating Tumor Cells in Unresectable Pancreatic Ductal Adenocarcinomas

**DOI:** 10.1101/2021.04.29.21256283

**Authors:** Hyemin Kim, Chan Mi Heo, Jinmyeong Oh, Eun Mi Lee, Juhee Park, Se-Hoon Lee, Kwang Hyuck Lee, Kyu Taek Lee, Jong Kyun Lee, Yoon-Kyoung Cho, Joo Kyung Park

## Abstract

**Background:** Circulating tumour cells (CTCs) have emerged as liquid biopsy biomarker providing non-invasive assessment of cancer progression and biology. We investigated whether longitudinal analysis of CTCs could monitor disease progression, response to chemotherapy, and survival in patients with unresectable pancreatic ductal adenocarcinoma (PDAC).

**Methods:** CTCs were isolated using a centrifugal microfluidic disc from serially collected peripheral blood with clinical assessments. CTCs were enumerated with immunostaining against Epithelial cell adhesion molecule, Cytokeratin, Plectin-1 and CD45.

**Results:** CTCs were detected in 91.7% of 52 patients with unresectable PDAC at the time of diagnosis. CTC numbers were not statistically different across tumour sizes, stages and metastatic sites. The absolute CTC counts after chemotherapy was inversely related to survival, and the decreased number of CTCs after the first cycle of chemotherapy was significantly associated with longer survival.

**Conclusions:** Identifying CTCs and monitoring CTC changes after chemotherapy could be a useful prognostic marker for survivals in patients with unresectable PDACs.

**Funding:** This work was supported by a grant from SK Chemical Research Fund of the Korean Society of Gastroenterology (Grant No.800-20130378) and a grant from Korean Gastroenterology Fund for Future Development. This study was granted by the Korean Health Technology R&D Project, Ministry of Health & Welfare funded by the Korean Government (Grant No. HI12C1845), and work by Y.K.Cho was partially supported by IBS-R020-D1 funded by the Korean Government. This research was supported by Collaborative Genome Program for Fostering New Post-Genome industry through the National Research Foundation (NRF) funded by the Korean government (MSIT) (Grant No. NRF-2017M3C9A5031002), and also supported by National Research Foundation (NRF) grant funded by the Korean government (MSIT) (Grant No. 2019R1C1C1008646).

**Clinical Trial Number:** ClinicalTrials.gov Identifier No. NCT02934984

## INTRODUCTION

Pancreatic ductal adenocarcinoma (PDAC) is the most common cancer with lethal effects, and the overwhelming majority of patients with PDAC have a locally advanced or distant metastatic disease (80–85%) (1, 2). It is currently the fourth leading cause of cancer-related deaths in the United States and is projected to be the second most common cause by 2030 (3). Despite many efforts to improve survival, it is still a lethal disease with a 5-year survival rate of less than 5% and a median survival of less than 1 year (4). The reasons for such poor survival are the lack of symptoms and effective ways to screen for pancreatic cancer, which result in a delayed detection of cancer (5). The computed tomography (CT) scan and endoscopic ultrasound (EUS)-guided biopsy become classic tool for the diagnosis with pathology confirmation of unresectable PDACs. Thus, there are clinical unmet needs for biomarker either tissue based or liquid biopsy to monitor and predict its prognosis in unresectable PDACs, especially. EUS-guided fine needle aspiration/biopsy (FNA/B) is still considered an invasive procedure; hence, it is unfit to use for monitoring disease progression and detecting molecular genetic changes during the courses of treatment. There have been many investigations to find biomarkers for PDAC and CA19-9 is, so far, the only biomarker with somewhat clinical usefulness. It is used for therapeutic monitoring and early detection of recurrent disease after treatment in pancreatic cancer (6, 7). However, it is not a specific biomarker to pancreatic cancer; CA19-9 level is also elevated in other conditions like cholestasis, lung diseases and other malignancy as well. In addition, approximately 10% of patients with PDACs who are negative for Lewis antigen a or b cannot synthesize CA19-9 (8).

Circulating tumour cells (CTCs) are rare, viable, tumour-derived epithelial cells identified in the peripheral blood of patients with cancer, which accompany tumour invasion into the blood. The ability to detect and analyse these CTCs in PDAC may give us insights into its aggressive biology (9, 10). Several studies have focused on identifying CTCs in the blood for diagnosis, staging and prognostication for various cancers (11-13). There have been studies in other tumour models, such as lung, breast, and prostate cancers that suggest that the presence of CTCs in the peripheral circulation of patients with metastatic carcinoma is associated with shorter survival (14-16). Unfortunately, there are only a few studies about the detection of CTCs in PDAC.

Multiple strategies for CTC isolation and identification have been reported (17-19); however, there have been emerging problems due to the extreme rarity, short lifetime and heterogeneity of CTCs. For example, antigen-dependent capture using an epithelial marker, Epithelial cell adhesion molecule (EpCAM), is a common isolation method, which can overlook CTCs undergoing an epithelial-to-mesenchymal transition (EMT) (20). Therefore, we additionally utilised a PDAC-specific marker, Plectin-1, to detect CTCs from patients with PDAC. Plectin-1 is known as a novel biomarker for primary and metastatic PDAC. It was identified in 100% of invasive PDAC tumours and 60% of pre-invasive Pancreatic intraepithelial neoplasia (PanIN) III lesions and was retained in metastatic deposits. Moreover, Plectin-1 distinguished PDAC from benign inflammatory diseases, like chronic pancreatitis (21). Recently, we have reported that captured PDAC CTCs were confirmed using Plectin-1 detection antibody as well as EpCAM antibody with KRAS mutation (22). Here, we examined whether Plectin-1 combined with EpCAM could improve CTC detection and the Plectin-1 positive PDAC CTCs can be used as prognostic biomarkers. The aims of this study are i) to enumerate CTCs using EpCAM and Plectin-1 antibodies in the peripheral circulation of patients with unresectable PDACs and ii) to investigate the clinical significance of CTC profiles correlated with clinical outcomes, such as treatment response, OS and site of metastasis in PDAC.

## RESULTS

### EpCAM, CK and Plectin-1 for CTC detection

EpCAM and CK are often overexpressed by epithelial carcinoma cells such as lung, colorectal, breast, prostate, head and neck, and liver, but absent in hematologic cells. Therefore, typical CTC enumeration method relies on the positive immunostaining of EpCAM and CK. While EpCAM has been widely used for immunospecific capture of CTCs from unfractionated blood, it cannot capture the CTCs under the epithelial-mesenchymal transition (20). We first confirmed the expression of EpCAM and CK in human pancreatic cancer cell lines by flow cytometry to determine if these antigens would be suitable for enumeration of PDAC CTCs. Pancreatic cancer cell lines, except PANC1 and Mia-PaCa2, abundantly expressed EpCAM on their surfaces (**Figure 1A**). In addition, all pancreatic cancer cell lines expressed the CK antigens, mostly at very high levels (**Figure 1B**). In these EpCAM/CK-expressing cells, these antigens were uniformly expressed throughout the entire population whereas WBCs did not express these antigens (**Figure 1A and B**).

**Figure 1.**
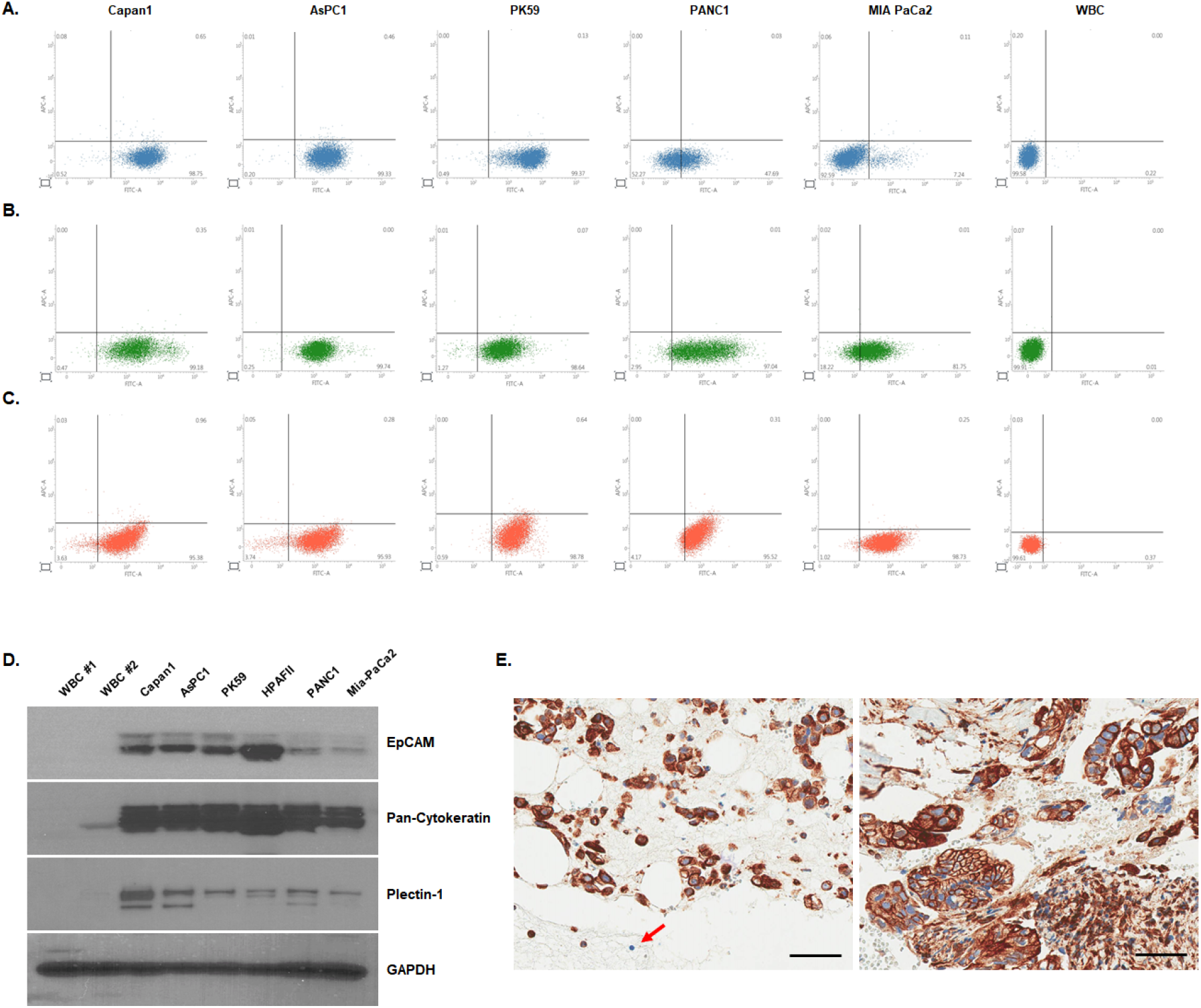
The expression EpCAM, Cytokeratin and Plectin-1 in PDAC cells. To validate EpCAM, Cytokeratin (CK) and Plectin-1 antibodies for detecting CTCs, the expression of EpCAM, CK and Plectin-1 antigen was examined in PDAC cell lines. The surface expression of (A) EpCAM and the intracellular expression of (B) pan CK and (C) Plectin-1 in PDAC cell lines were examined by flow cytometry. WBCs were used as negative controls. (D) The expression of EpCAM, pan CK, and Plectin-1 in PDAC cell lines were determined by immunoblotting, and (E) the expression of Plectin-1 in PDAC tissues were evaluated by immunohistochemistry. Arrow indicates a WBC. Scale bar, 50 μm.

In addition, we tested Plectin-1, a novel PDAC-specific biomarker reported to be expressed on the membrane and cytoplasm of PDAC cells (21). We could find out that all PDAC cell lines expressed the Plectin-1 antigen and it was uniformly expressed throughout the entire population, whereas WBCs rarely express this antigen (**Figure 1C**). Moreover, immunoblot analysis showed the expression of EpCAM, CK and Plectin-1 in pancreatic cancer cells but not in WBCs (**Figure 1D**). Also, we observed an intensive expression of Plectin-1 in ductal cancer cells of the PDAC tissue except WBCs through IHC staining (**Figure 1E**). Plectin-1 was thoroughly expressed in the metastatic liver tissue from PDAC patients as well, whereas its expression was very low in normal pancreas tissue and immune cells in palatine tonsil or intestine tissue (**Supplementary Figure 1**). We previously reported that healthy subjects had negligible CTC counts (mean, 0 CTCs/7.5 mL; median, 0 CTCs/7.5 mL; range, 0-5 CTCs/7.5mL of blood) when EpCAM and CK antibodies were used to detect CTCs (23, 24). Recently, we have also showed the specificity of Plectin-1 to capture CTCs in PDAC (22). In twenty-eight blood samples (3ml/each) from healthy donors, there was no single positive staining of Plectin-1 for CTCs in 17 samples (61%), and 11 healthy donors (39%) had 0.3∼0.7 CTCs per ml of blood (Mean±SD, 0.16±0.21 CTC/ml; Median 0.00 CTC/ml) (**Supplementary Figure 2**).

### Enrichment and identification of CTCs in patients with PDAC

PDAC CTCs were enriched using ‘FAST disc’, a centrifugal microfluidic tangential flow filtration device, which allowed rapid, label-free isolation of CTCs from whole blood without sample pre-treatment (24). First, the membrane of FAST disc was examined after running the blood samples of PDAC patients by haematoxylin and eosin (H&E) staining (**Figure 2A**). We found CTCs captured on the membrane, which was further confirmed as cancer cells by specialized pathologists.

**Figure 2.**
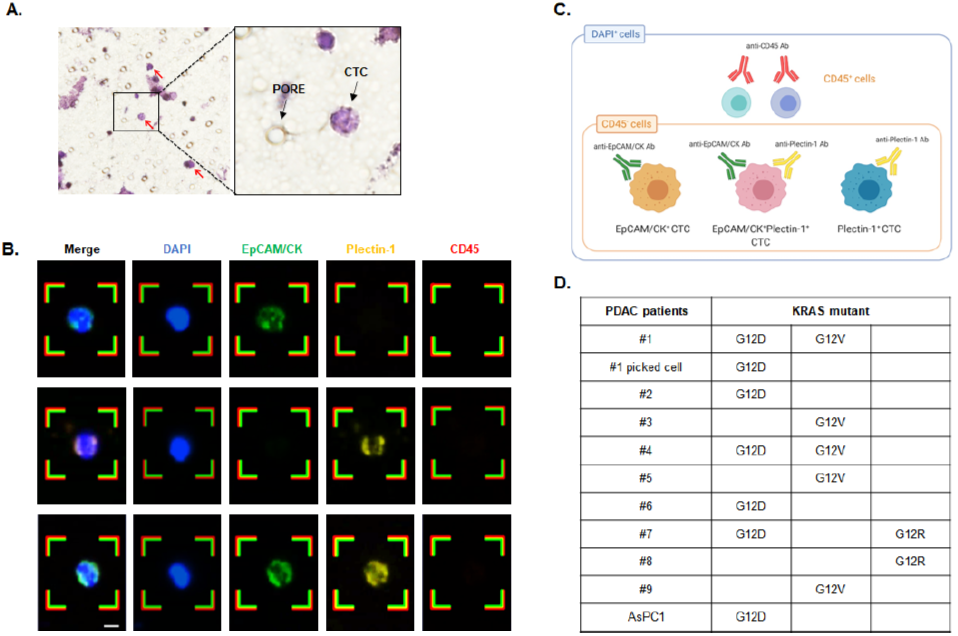
Identification of CTCs in patients with PDAC. PDAC CTCs were identified by haematoxylin and eosin (H&E) and immunofluorescent (IF) staining. (A) Captured CTCs on the membrane of a disc were pathologically identified by H&E staining. Arrows indicate captured CTCs. Membrane pore size is 8 μm. (B) Three representative images of PDAC CTCs. All nucleated cells were stained by DAPI (Blue), WBCs were identified by CD45 (Red) staining, and CTCs were identified by EpCAM/CK (Green) and Plectin-1 (Gold) staining. Scale bar, 5 μm. (C) PDAC CTCs were defined as the sum of EpCAM/CK^+^, Plectin-1^+^, and EpCAM/CK/Plectin-1^+^ cells among DAPI^+^ and CD45^-^ cells. Illustration was created with Biorender.com. (D) Digital droplet PCR for 3 types of KRAS mutants were performed to confirm PDAC CTCs captured on the membrane of the FAST disc. AsPC1 was used for a positive control.

In addition, PDAC CTCs were identified by immunofluorescence (IF) staining with anti-EpCAM/CK, anti-CD45 and anti-Plectin-1 antibodies (**Figure 2B**). Among DAPI^+^ and CD45^-^ cells, we could find three kinds of CTCs: EpCAM/CK positive (ranged from 0 to 631 cells/3 ml), Plectin-1 positive (ranged from 0 to 123 cells/3 ml), and both EpCAM/CK and Plectin-1 positive (ranged from 0 to 127 cells/3 ml) cells. We decided to consider all three cases (ranged from 0 to 641 cells/3 ml) as PDAC CTCs (**Figure 2C**).

Next, we performed molecular characterization of the cells captured on the membrane of the FAST disc. KRAS mutation is the most frequently detected somatic alteration in PDACs (nearly 100%) (25) and thus the detection of KRAS mutants can be a proof to confirm that the captured cells are CTCs of PDACs. After the enrichment of CTCs from the blood samples of patients with PDAC, DNAs were extracted from the cells captured on the membrane and followed by ddPCR. Three types of KRAS mutant (G12D, G12V, G12R) were detected in CTCs from patients with PDAC (**Figure 2D** and **Supplementary Figure 3A**). AsPC-1 was used as a positive control for PDAC, which had KRAS G12D mutant. Moreover, we picked single Plectin-1^+^ cell (**Supplementary Figure 3B**) and confirmed KRAS G12D mutant (a single CTC isolated from patient #1), which was also detected from the sample prepared from the whole membrane capturing CTCs (**Figure 2D**).

### Baseline characteristics of study patients

A total of 52 patients were enrolled in this study, and their baseline characteristics are described in **Table 1**. There were 26 male and 26 female patients aged 46–85 years (median 67 years). Clinical stage (8^th^ AJCC) was as follows: 21 were stage III (40.4%) and 31 were stage IV (59.6%). Twenty patients (38.5%) had metastasis to the liver, and 11 patients (21.1%) showed metastasis to bone, peritoneum and supraclavicular lymph node (SCN) without liver. The 46 patients (88.5%) received chemotherapy and 6 patients (11.5%) did not receive chemotherapy due to patient preference, old age or poor performance status. Median (range) of initial CA19-9 and CEA were 203.5 IU/ml (3.75–140000 IU/ml) and 2.9 ng/mL (0.5–51.39 ng/mL), respectively.

**Table 1.** Patient characteristics (n = 52)

| Characteristics | n = 52 |
| --- | --- |
| Age, median (range) | 67 (46-85) |
| Sex, n (%) |  |
| Male | 26 (50.0) |
| Female | 26 (50.0) |
| BMI (kg/m<sup>2</sup>), median (range) | 23.6 (19.0-34.1) |
| Performance status (ECOG), n (%) |  |
| 0: fully active | 28 (53.9) |
| 1: light house work | 22 (42.3) |
| 2: ambulatory | 1 (1.9) |
| 3: limited self-care | 1 (1.9) |
| Stage (AJCC 8<sup>th</sup>), n (%) |  |
| III | 21 (40.4) |
| IV | 31 (59.6) |
| Metastasis, n (%) |  |
| No metastasis | 21 (40.4) |
| Liver metastasis | 20 (38.5) |
| Other site metastasis, not including liver | 11 (21.1) |
| Location (proximal), n (%) |  |
| Uncinate/head/Neck | 24 (46.1) |
| Body | 16 (30.8) |
| Tail | 12 (23.1) |
| Pancreas mass (mm), median (range) | 35 (14.0-130.0) |
| Chemotherapy |  |
| No chemotherapy | 6 (11.5) |
| Gemcitabine based | 24 (46.2) |
| FOLFIRINOX | 21 (40.4) |
| TS-1 | 1 (1.9) |
| CA19-9 (IU/mL), median (range) | 203.5 (3.75-140000) |
| CEA (ng/mL), median (range) | 2.9 (0.5-51.39) |

BMI, Body mass index; ECOG, Eastern Cooperative Oncology Group; AJCC, American Joint Committee on Cancer; CA19-9, carbohydrate antigen 19-9; CEA, Carcinoembryonic antigen.

### Evaluation of CTCs from the study patients

Fifty-two blood samples from patients with PDAC were analysed at baseline. The median number of CTCs was 26 (range 0-641 cells/3 ml), and 91.7% of patients with PDAC had one or more CTC at baseline. We evaluated the tumour size, stage and metastatic sites based on the number of CTCs to demonstrate the unique clinical potential of CTCs. We calculated the sum of the unidimensional size in centimetres of all significant and measurable tumour sites through the CT scan of each patient. The absolute number of CTCs captured did not necessarily correspond with tumour size (tumour median size, 36 mm; p=0.5139, **Figure 3A**). In addition, CTC counts were remarkably higher in stage III than IV (*p=0.0403, **Figure 3B**). The most common site of metastasis is the liver in PDAC due to the fact that the first venous drainage of pancreatic cancer is the portal circulation. There was no significant difference in the number of CTCs across the metastatic sites (p=0.077), but patients having multi-metastases to the liver and other sites, such as the peritoneum, lung, bone and SCN had increased CTC counts (**Figure 3C**).

**Figure 3.**
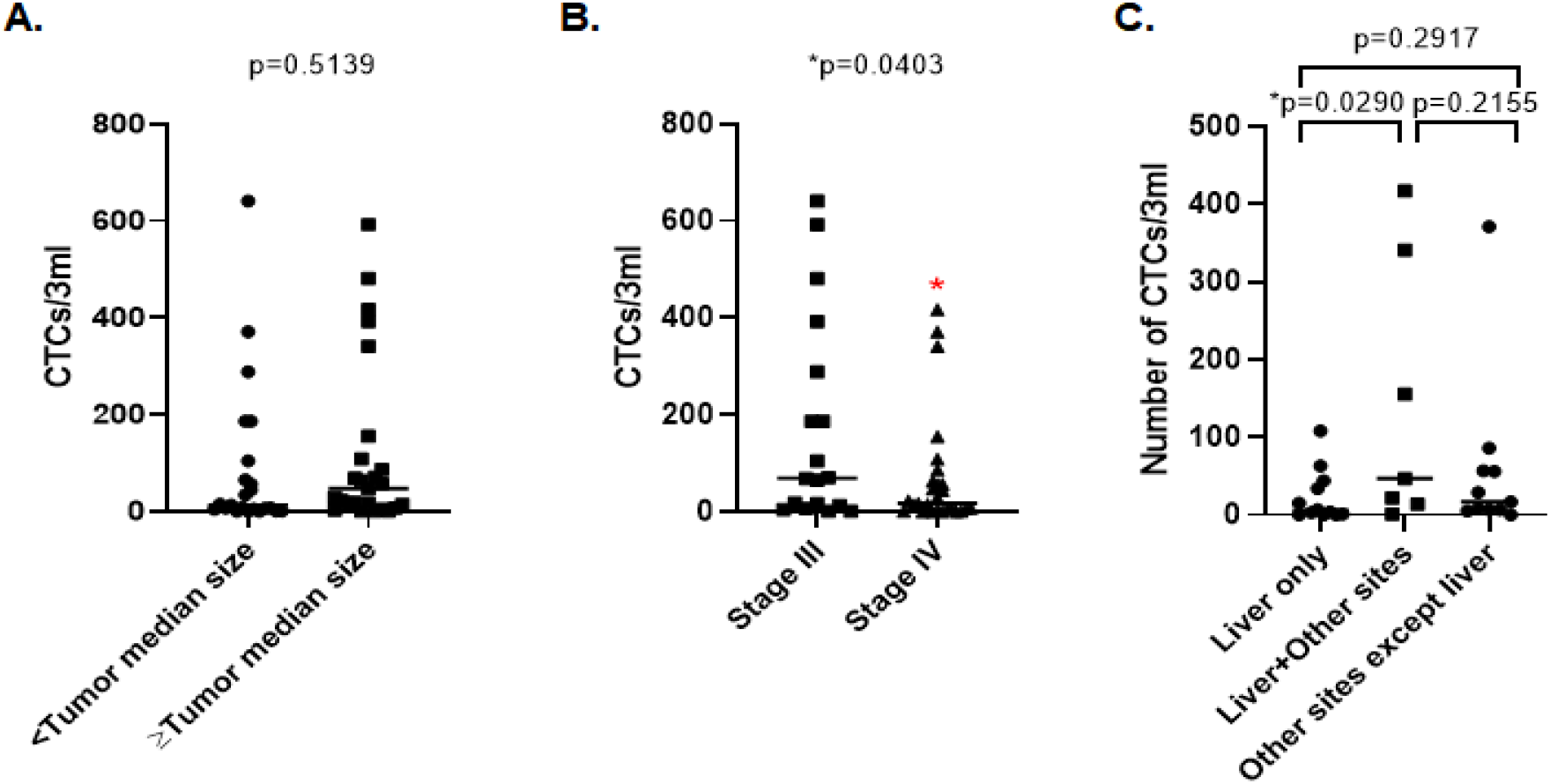
CTC enumeration in patients with PDAC. CTCs were isolated and counted in blood samples from 52 patients with PDAC. CTC counts were analysed depending on (A) tumour size (median 36 mm, p=0.5139), (B) clinical stages (*p=0.0403) and (C) metastatic sites (p=0.077). Other sites include lung, bone, supraclavicular lymph node (SCN) and peritoneum. Each bar represents the median.

### Assessment of CTC numbers before and after chemotherapy

CTCs were evaluated in patients who underwent chemotherapy (CTx). Blood samples were collected at baseline (pre-CTx) and during subsequent clinic visits for treatment (post-CTx). We could collect full series of blood draws both pre-CTx and post-CTx from thirty-nine patients according to the performance status. The exact follow-up schedule varied between patients. CT scan and serum CA19-9 measurement were performed at baseline and at regular intervals according to standard clinical practice (**Figure 4A**). Upon comparing CTC counts in pre- and post-CTx blood samples of patients with PDAC, we found that the absolute number of CTCs was not remarkably decreased after treatment (p=0.5496, **Figure 4B**). There was no significant difference in CTC counts at baseline according to tumour location (p=0.3970, **Figure 4C**); however, CTC counts were higher in head PDAC than body/tail PDAC after treatment (*p=0.0253, **Figure 4D**).

**Figure 4.**
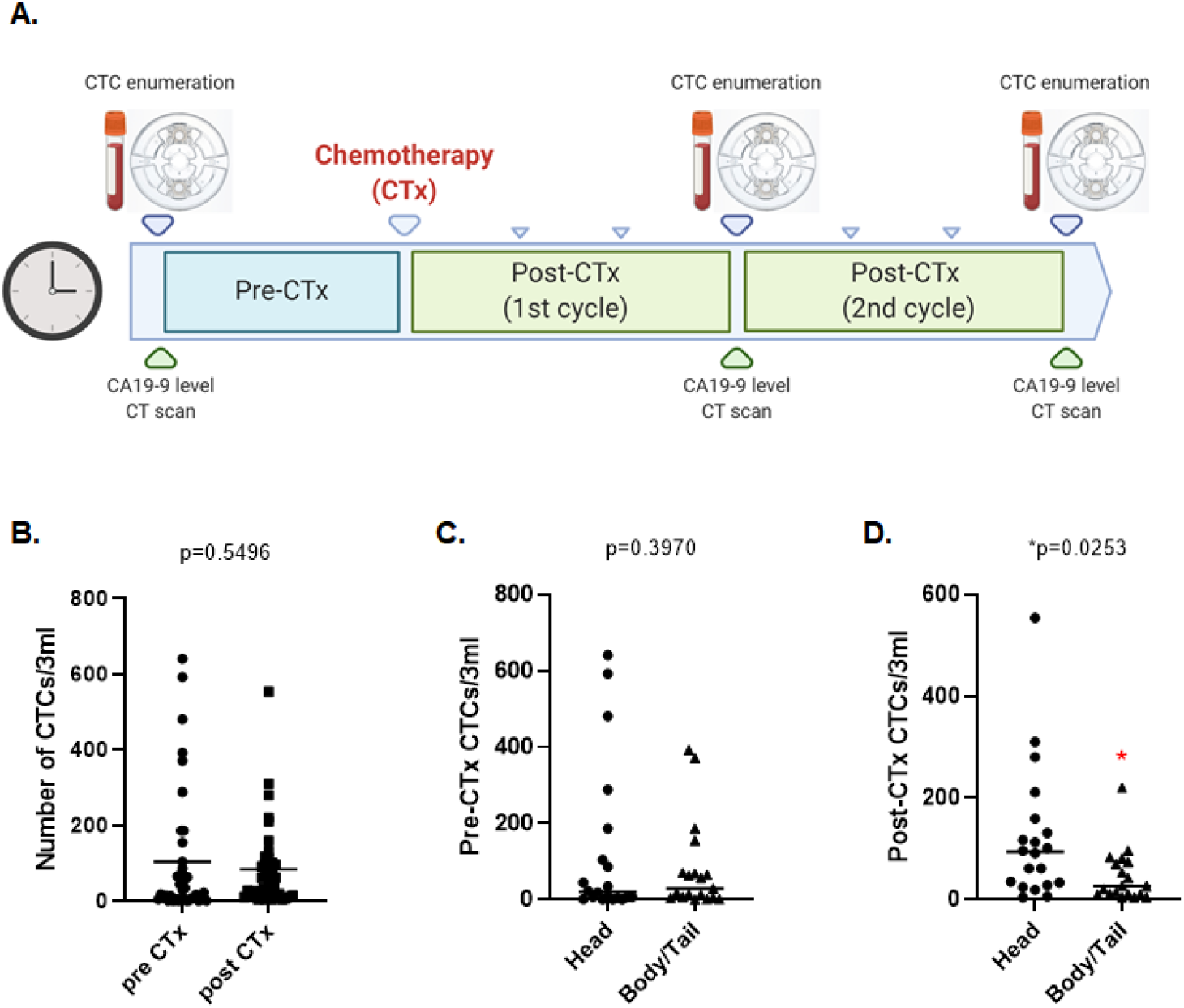
Evaluation of CTC counts in PDAC patients with chemotherapy. (A) CTCs were enumerated from paired blood samples of 39 patients with PDAC before (Pre-CTx) and after chemotherapy (post-CTx), which was accompanied with measurement of serum CA19-9 level and CT scanning. Illustration was created with BioRender.com. (B) The absolute number of CTCs at the pre-CTx and post-CTx was evaluated (p=0.5496). The number of CTCs at (C) Pre-CTx and (D) Post-CTx was analysed with tumour locations divided by head and body/tail (p=0.3970 and *p=0.0253, respectively). Each bar represents the median. CT, computed tomography; CA19-9, carbohydrate antigen 19-9.

In addition, absolute CTC counts at post-CTx (*p=0.0471) could predict survival of patients with PDAC in which the number of CTCs over the median (47.5/3 ml blood) at post-CTx was associated with worse prognosis. However, this correlation was not observed with pre-CTx counts (p=0.3088) (**Figure 5A** and **5B**).

**Figure 5.**
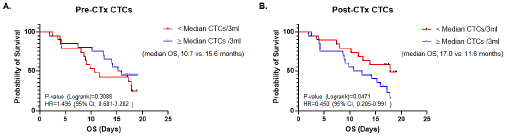
Kaplan–Meier analysis with the number of CTCs and overall survival in patients with PDAC with chemotherapy. Kaplan–Meier curves of overall survival (OS) and log-rank tests for patients with PDAC depending on the number of CTCs (A) before chemotherapy (pre-CTx) and (B) after chemotherapy (post-CTx). No statistical difference in OS was observed at pre-CTx (p=0.3088), but the number of CTCs at post-CTx was related to OS of patients with PDAC (*p=0.0471).

### Clinical significance of CTC changes after chemotherapy

We calculated the change in CTC numbers after chemotherapy relative to the number of CTCs at baseline, and we evaluated its relationship with chemotherapy responses and OS of patients with PDAC. A higher CTC count at post-CTx compared to pre-CTx was more evident in patients who poorly responded to the treatment (stable disease [SD]/progressive disease [PD]) than in patients who favourably responded to the treatment (complete remission [CR]/partial regression [PR]), although there was no statistical significance (**Figure 6A**). Absolute numbers of CTC were significantly elevated in the increased CTC group of patients with PDAC after chemotherapy (*p=0.0208, **Figure 6B**). Furthermore, the Kaplan–Meier analysis indicated that patients who had relatively increased CTCs after chemotherapy showed worse prognosis (**Figure 6C**). The OS was longer when the number of CTCs were reduced after chemotherapy, and patients with PDAC with an increase of CTCs showed shorter survival (median OS, 16.97 vs. 10.02 months; **p=0.0095). The association of CA19-9 changes with patient survival was also examined as CA19-9 is the most commonly used biomarker for diagnosis and management of patients with pancreatic cancer (26). The increase or decrease of CA19-9 level with chemotherapy failed to prognose the survival of patients with PDAC (**Figure 6D**).

**Figure 6.**
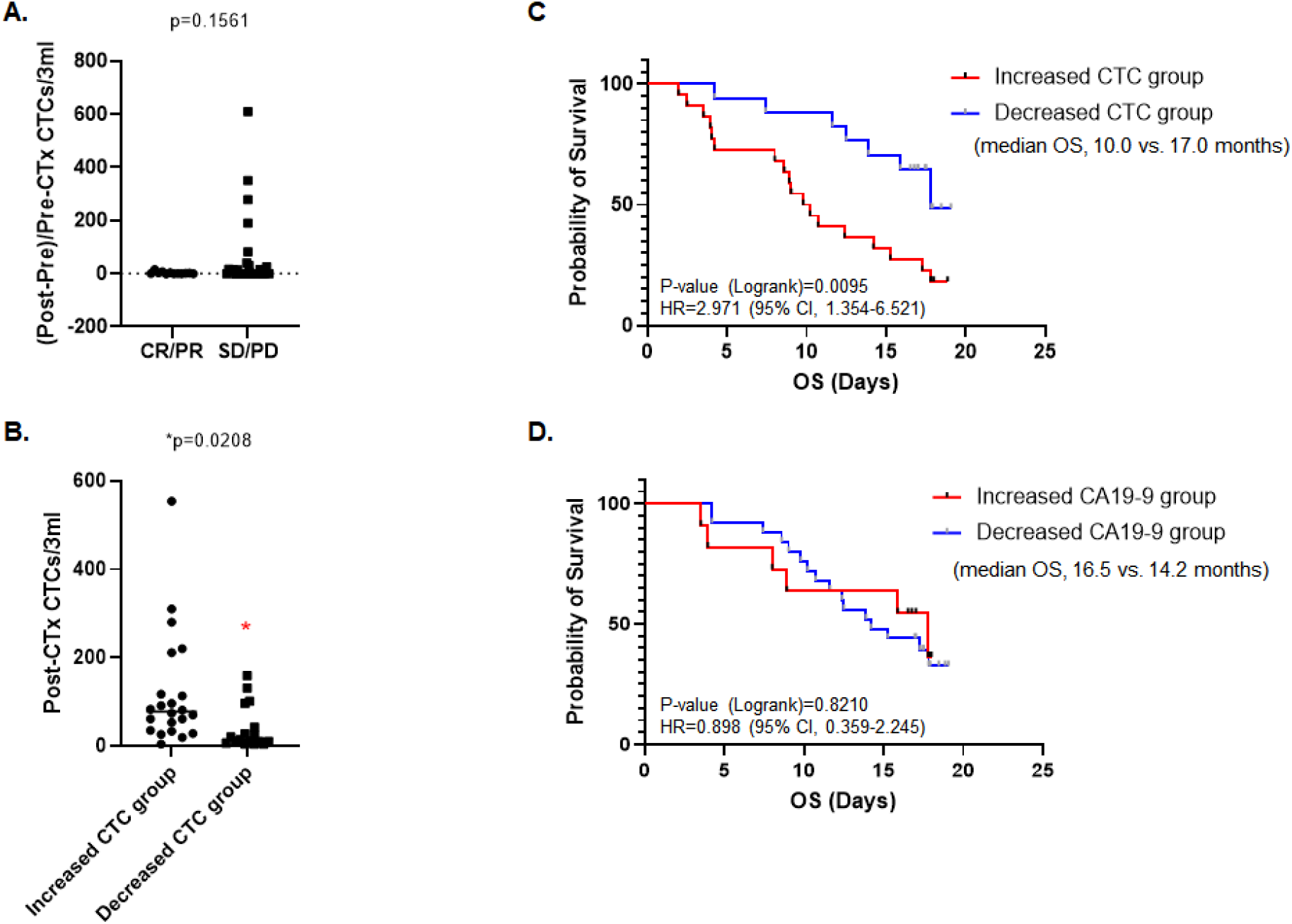
Evaluation of CTC changes after chemotherapy in patients with PDAC. The change of CTC numbers after chemotherapy (post-CTx) relative to before chemotherapy (pre-CTx) was evaluated with chemotherapy responses and overall survival (OS) of patients with PDAC. (A) The relative change of CTC counts was assessed with responses to chemotherapy in patients with PDAC. There was no significance between the completely or partially responded (CR/PR) group and stably or progressively responded (SD/PD) group (p=0.1561). (B) The number of CTCs was compared between patients with increased CTCs (increased CTC group) and patients with decreased CTCs (decreased CTC group) after chemotherapy (*p=0.0208). Each bar represents the median. Kaplan–Meier survival curve was stratified by the comparative changes (increased or decreased) of (C) CTC counts and (D) serum CA19-9 levels after chemotherapy. Significance was observed in the change of CTC numbers (**p=0.0095), but not in the change of CA19-9 levels (P=0.8210).

## DISCUSSION

Investigation into CTCs may give us insights on the biology of tumour cell dissemination in patients with cancer. Little is known about the biology and pathology of CTCs in PDAC due to difficulty in collecting them in the peripheral circulation. Here we efficiently collected CTCs from blood samples of patients with PDAC by using size-based centrifugal microfluidic disc and by using both EpCAM, CK, and Plectin-1 identification antibodies to estimate their prognostic potential for responses to treatment and survival.

Tumour cells in the peripheral blood are substantially heterogeneous, which offers a unique opportunity to understand how CTCs participate in the tumour dissemination process and tumour heterogeneity (10). However, these heterogeneous properties of rare CTCs make it harder to detect their variable phenotypes in circulation. The only system currently approved by the Food and Drug Administration (FDA) as an aid in monitoring patients with metastatic breast, colorectal or prostate cancer is CELLSEARCH® (Janssen Diagnostics, Raritan, NJ, UA), which uses antibodies specific to EpCAM and CK of epithelial CTCs (18). However, CTCs exhibit dynamic changes in epithelial and mesenchymal compositions (27), and show both epithelial and mesenchymal features (28). They can show no or low expression of EpCAM during EMT, resulting in missed detection (29). Furthermore, EpCAM is down-regulated in pancreatic cancer (30, 31). To make up for this, we decided to use a PDAC-specific antibody in addition to EpCAM and CK. The ideal biomarker for PDAC should not only differentiate benign conditions from malignancy but also be able to detect small cancers, ideally at the pre-invasive PanIN III phase. Plectin-1 expression is positive in all PDACs but negative in benign tissues. Moreover, it could detect pre-invasive PanIN III lesions (21). Therefore, we additionally utilised a PDAC biomarker, Plectin-1, to identify CTCs in the peripheral blood samples of patients with PDAC. As a result, we were able to successfully detect cells positive to only Plectin-1 and not EpCAM/CK in the bloodstream (**Figure 2B**). Moreover, KRAS mutant was detected in both Plectin-1^+^ cells and CTCs captured on the membrane (**Figure 2D**). Thus, PDAC CTCs were defined as all EpCAM/CK orPlectin-1-positive cells in our study (**Figure 2C**). CTC enumeration with an additional Plectin-1 antibody could more significantly predict OS compared to CTC enumeration with only EpCAM/CK antibodies (**p=0.0095 vs. *p=0.0152, HR=2.971 vs. HR=2.689, **Figure 6C** and **Supplementary Figure 4**).

The enumeration of CTCs was not statistically different in relation to tumour size and metastatic sites (**Figure 3A** and **B**). Patients with stage III PDAC showed more CTCs than patients with stage IV PDAC (**Figure 3B**). CTCs mediate the metastatic dissemination of cancer (32), and can be expected to include the subpopulations responsible for disease progression (10). Seventy-three percentage of patients with stage III PDAC showed poor responses, and it is likely that the high number of CTCs could mean the potential of progression. It is worth further investigation.

Our study was designed such that CTCs were identified both before and after chemotherapy with the clinical assessment of the CT scan and serum CA19-9 measurement (**Figure 4A**). The number of CTCs was not significantly decreased by chemotherapy across all patients with PDAC (**Figure 4B**). The detection and existence of CTCs can be a key model of haematogenous spread in the development of metastatic disease. The existence of CTCs expressing the cell surface EpCAM and intracellular CKs is related to poor outcome in patients with both non-metastatic and metastatic disease (33-35). In our result, patients with PDAC with a high number of CTCs showed worse prognosis, but CTC counts had prognostic value for survival prediction only at post-CTx and not at pre-CTx (**Figure 5A** and **5B**). Furthermore, the relative change between pre-CTx and post-CTx has more critical prognostic significance to predict the probability of survival. Patients with decreased CTCs after chemotherapy indicated a significantly longer survival, whereas patients with increased CTCs showed poor survival (**Figure 6C** and **Supplementary Figure 5**). However, it seems that the evaluation of both absolute CTC counts and relative CTC changes with chemotherapy is not enough to predict chemotherapy responses (**Figure 6A**). Considering that the blood collection was only performed after the first cycle of the treatment, there is a need for further investigation to reveal the association of CTC counts with poor (SD/PD) or good (CR/PR) responses to chemotherapy.

The sensitivity of CTC capture and identification was 91.8% in our results, which is higher than other reports. Sefrioui *et al* reported 67% sensitivity of CTC in solid pancreatic tumours (36), Ankeny *et al* reported 75% sensitivity of peripheral CTC in pancreatic cancer and Kulemann *et al* reported 67.3% sensitivity in pancreatic cancer (37). Serum level of CA19-9 is the only marker approved by the United States FDA for use in the routine management of pancreatic cancer (38, 39). The sensitivity of CA19-9 is 63.6%, and changes in serum levels are unrelated to disease progression (30). It coincided with a result where an increased or decreased level of CA19-9 with chemotherapy was not related to the probability of survival (**Figure 6D**) as opposed to significant correlation of CTC changes with survival (**Figure 6C**).

The direction of systemic cancer treatment based on the primary tumour characteristics has limitations due to the tumour heterogeneity and frequent discrepancy between primary and metastatic sites. However, because of both inaccessibility of metastatic sites and procedure morbidity, metastatic biopsies are rarely undertaken (40, 41). In this sense, the prognostic role of CTC enumeration is the true promise to provide a real-time view of cancer progression just using peripheral blood samples, avoiding the need for repeat invasive biopsies. Understanding the biology of CTCs or cancer cells in transit may give us unique insights into the mechanisms behind metastasis. In addition, further genomic analysis of CTCs needs to be performed.

## MATERIALS AND METHODS

### Validation of Capture and Identification Antibodies

Pancreatic cancer cell lines were maintained as a monolayer in culture media including Dulbecco’s Modified Eagle’s Medium for PANC1 and MIA PaCa2 cells, RPMI-1640 Medium for AsPC1 and PK59 cells, and Iscove’s Modified Dulbecco’s Medium for Capan1 cells at 37°C in a humidified atmosphere containing 5% CO2. Pancreatic cancer cells were detached with TrypLE™ Express Enzyme solution (ThermoFisher Scientific, Rockford, IL, USA) and washed with phosphate buffered saline (PBS). For flow cytometry, white blood cells (WBCs) and harvested pancreatic cancer cells (1⨯10^6^ cells/tube) were incubated with FITC-conjugated anti-EpCAM (ThermoFisher Scientific) for cell surface staining. After washing with Bovine serum albumin (BSA, 0.2%) stain buffer (BD Biosciences, San Jose, CA, USA), cells were fixed and permeabilized with Cytofix/Cytoperm™ Fixation/Permeablization solution (BD Biosciences) for 30 min at room temperature (RT). After washing with Perm/Wash buffer (BD Biosciences), resuspended cells were incubated with Alexa Fluor 488-conjugated Pan Cytokeratin (CK) Monoclonal Antibody (AE1/AE3) (ThermoFisher Scientific) or Plectin-1 Monoclonal Antibody (D6A11) (Cell Signaling Technology (CST), Danvers, MA, USA) for 1 hour on ice with gentle shaking in the dark. Followed by anti-rabbit IgG (H+L), F(ab’)2 Fragment (Alexa Fluor® 488 Conjugate) (CST) incubation for Plectin-1, washed cells were analysed on BD FACSVerse™ flow cytometer and BD FACSuite™ Software (BD Biosciences). For immunoblot of PDAC cell lines and WBCs, proteins (30 μg) were separated by using NuPAGE™ Tris-Acetate (3-8%) gel electrophoresis system (ThermoFisher Scientific), and the expression of EpCAM, Pan CK, Plectin-1 and GAPDH was detected by Pierce™ Enhanced chemiluminescence system (ThermoFisher Scientific). For immunohistochemistry (IHC) of PDAC tissues, paraffin-embedded sections (5 μm) were deparaffinized and hydrated. Followed by epitope retrieval with BOND ER1 buffer (pH6.0, Leica Biosystem, Melbourne, Australia) at 100°C, primary Plectin-1 antibody (1:300, Abcam, Cambridge, UK) was incubated for 15 min, and Bond™ Polymer refine detection (Vision Biosystems, Melbourne, Australia) for 10 min with Bond-Rx autoimmunostainer (Leica Biosystem). After counterstaining with hematoxylin and mounting, stained slides were scanned with ScanScope® AT System (Aperio Technologies, Vista, CA, USA).

### Study Patients and Blood collection

Fifty-two patients with PDAC diagnosed between January and June 2019 at Samsung Medical Center were prospectively enrolled (ClinicalTrials.gov Identifier No. NCT02934984) and followed up until the end of 2020. Clinical Records Form (CRF) was prospectively collected and the following medical information was contained: age, sex, staging (The 8^th^ edition American Joint Committee on Cancer (AJCC)), body mass index, Eastern Cooperative Oncology Group (ECOG) performance status, comorbidity, serum levels of Carcinoembryonic antigen (CEA), carbohydrate antigen (CA) 19-9, chemotherapy regimen, clinical response and survival data. This study was conducted under the principles of the Declaration of Helsinki. The study protocol was approved by the institutional review board (IRB) of Samsung Medical Center. All patients provided written informed consent, and all specimens were collected according to IRB regulations and approval (IRB No. 2018-11-080). Blood was taken from the peripheral vein (cephalic vein) and collected into a Cell-Free DNA BCT® CE tube (Streck, Omaha, NE, USA). It was processed within 3–4 hours for CTC enrichment and enumeration.

### Microfluidic Approach to Isolate and Enumerate CTCs

The CD-PRIME™ system of the CD-CTC disposable disc and CD-CTC Enrichment kit (Clinomics, Inc., Ulsan, Korea) was used to isolate CTCs efficiently from millions of other blood cells in whole blood samples. This CTC enrichment system called ‘Fluid-Assisted Separation Technology (FAST) disc’ is based on centrifugal microfluidic separation (23, 24, 42). This device was previously reported by our group, and the real-time simulation was open to the public on YouTube (https://www.youtube.com/watch?v=OgJ8eztIYQA). It is based on the size-selective, clog-free CTC isolation through a polyethylene membrane (8-μm pore size) filled with a stably-held liquid throughout the filtration process. Briefly, the sample-loading chamber, filter zone and fluid assistant chamber were filled with a 1% bovine serum albumin (BSA) solution and rotated at a spin speed of 600 rpm for washing. Then, 3 ml of whole blood was loaded to the sample loading chamber and rotated at 600 rpm. After washing with the 1% BSA solution 3 times, CTCs captured on the disc membrane were fixed with 4% paraformaldehyde for 15 min at room temperature for the next CTC staining and enumeration processes.

### Staining and enumeration of CTCs

CTCs fixed on the membrane were immunostained using the CD-CTC cell enumeration kit (Clinomics, Inc., Ulsan, Korea) combined with Plectin-1 antibody to identify the number of isolated CTCs. The cells were permeabilised with 0.1% Triton x-100 in PBS for 5 min and blocked with IgG (20 ug/ml) or goat serum for 20 min after washing with PBS. Anti-Plectin-1 (1:60, CST) solution was applied at 4°C overnight. The next day, cells were additionally stained with Alexa 647-conjugated anti-rabbit IgG (1:500, CST), FITC-conjugated anti-CK (1:100), Alexa 488-conjugated anti-pan CK (1:100), FITC-conjugated anti-EpCAM (1:400) and Alexa 594-conjugated anti-CD45 (1:100). All antibodies, including pan CK, EpCAM and CD45 antibodies, were included in the cell enumeration kit. CTCs were identified by using an imaging system, consisting of staining with 4.6-diamidino-2-pheylindole (DAPI) for DNA content, fluorochrome-conjugated anti-CD45 for haematologic cells and anti-Plectin-1 or anti-EpCAM/CK for PDAC CTC cells. Then number of CTCs/ml was determined via comprehensive image analysis, scanning the entire membrane (Bioview CCBS system, BioView, Ltd., Nes Ziona, Israel) and identifying CTCs based on cell size, morphology and IF staining. Total number of cells was counted by DAPI staining; WBCs were identified by CD45 staining and capture of PDAC CTCs was confirmed by IF staining profiles: CD45 (WBC marker) negative and EpCAM/CK (epithelial marker) or Plectin-1 (PDAC-specific marker) positive cells. PDAC CTCs were defined as EpCAM/CK^+^CD45^-^DAPI^+^ cells, Plectin-1^+^CD45^-^DAPI^+^ cells and EpCAM/CK^+^Plectin-1^+^CD45^-^DAPI^+^ cells. Eleven blood samples (3 ml each) of control donors were analysed on the CTC discs to validate the specificity of EpCAM/CK and Plectin-1.

### Confirmation of KRAS Mutation in Captured CTCs

After CTC enrichment from the blood of PDAC patients, DNA was recovered from the captured CTCs on the FAST disc membrane with QIAamp DNA Micro Kit (Qiagen, Germantown, MD, USA). Mutant KRAS was detected by Droplet Digital polymerase chain reaction (ddPCR) to identify three somatic mutations located in codons 12 (p.Gly12Asp (G12D), p.Gly12Arg (G12R), p.Gly12Val (G12V)) in Macrogen Inc. (Seoul, Korea). Plectin-1^+^ CTCs on the membrane were picked by CellCelector (ALS Automated Lab Solutions GmbH, Jena, Germany), and whole genome amplification (WGA) was carried out using REPLI-g single cell kit (Qiagen). ddPCR was performed with 2X ddPCR Supermix and QX200 Droplet Digital PCR System (Bio-Rad, Hercules, CA, USA). Data analyses were performed as recommended by the manufacturer using the QuantaSoft Software version 1.7.4. (Bio-Rad).

### Statistics

Non-parametric tests were used throughout the study. The difference of CTC numbers was analysed by an unpaired t-test or one-way ANOVA. OS was analysed by the Kaplan–Meier method with the use of one-sided log-rank statistics. P values <0.05 were considered statistically significant. Statistical analysis was carried out using SPSS for Windows (version 17.0, SPSS Inc. Chicago, IL, USA) and GraphPad Prism 8.0 (GraphPad Software Inc., La Jolla, San Jose, CA, USA).

## Data Availability

All data generated or analysed during this study are included in the manuscript and supporting files.

## < Supplementary Figure Legends >

**Supplementary Figure 1.**
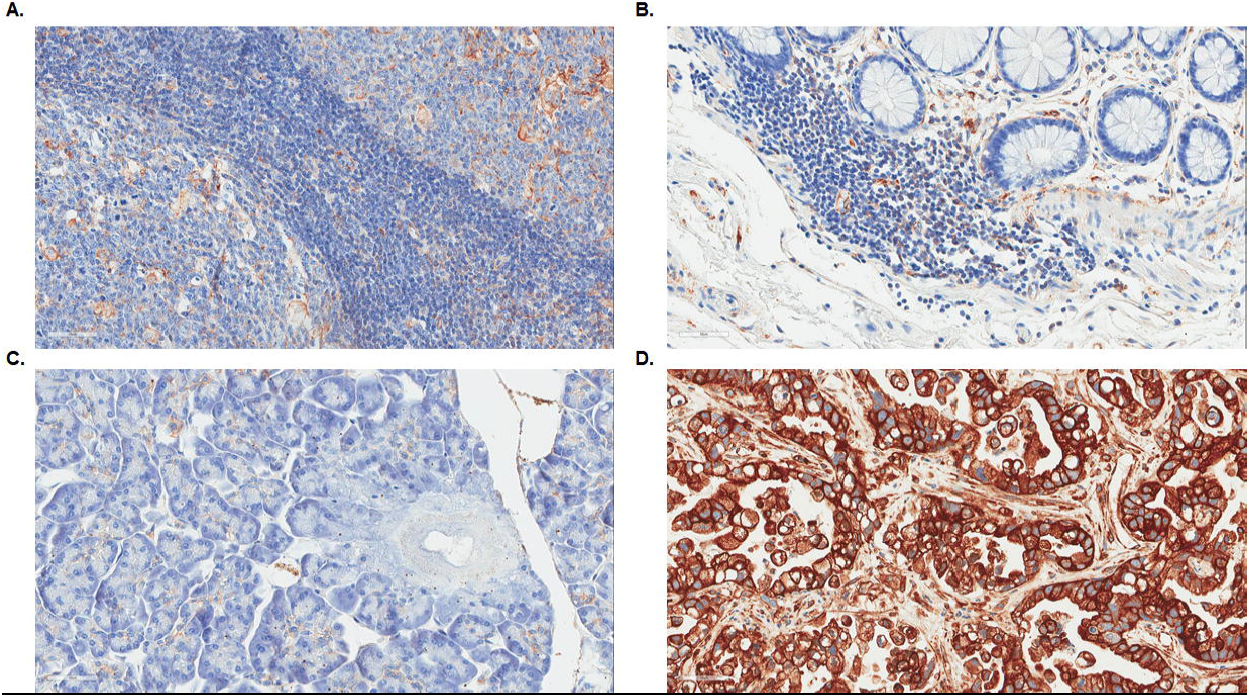
The expression of Plectin-1 in normal tissues and metastatic tissues. The expression of Plectin-1 in (A) tonsil, (B) colon, (C) normal pancreas, and (D) liver metastases from pancreatic ductal adenocarcinoma (PDAC) were examined with immunohistochemistry.

**Supplementary Figure 2.**
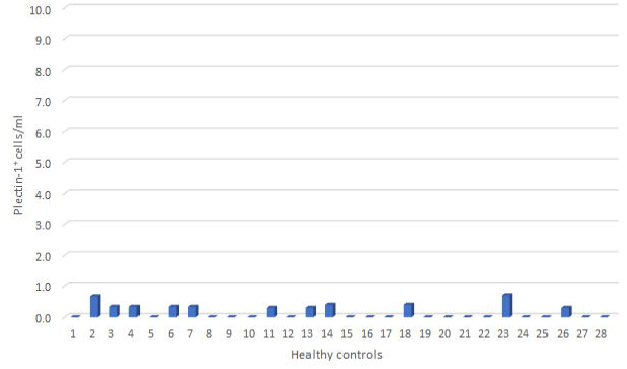
The number of Plectin-1+ CTCs in healthy controls. CTCs were isolated from 28 blood samples from healthy donors and enumerated with Plectin-1 staining. Plectin-1^+^ CTCs were represented as cells per 1ml blood for each individual.

**Supplementary Figure 3.**
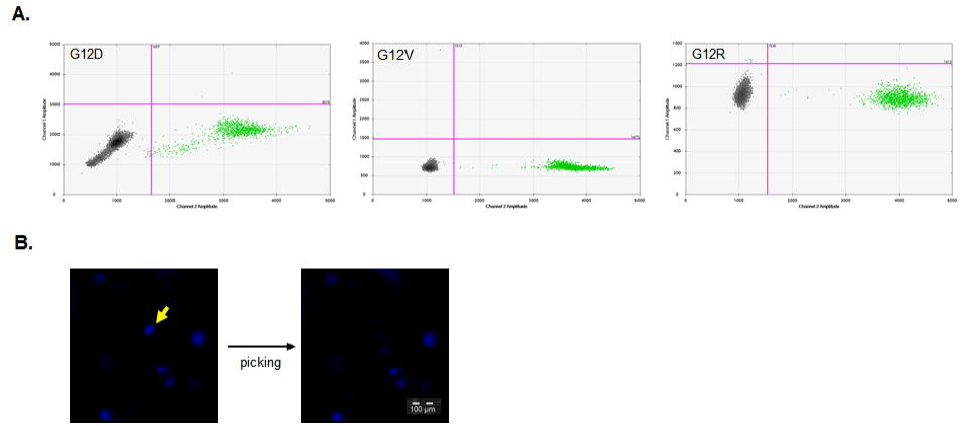
Digital droplet PCR (ddPCR) for the detection of KRAS mutants in CTCs isolated from PDAC patients. (A) Digital droplet PCR for 3 types of KRAS mutants were performed to confirm PDAC CTCs captured on the membrane of the FAST disc. (B) A single cell was picked from the membrane after staining with Plectin-1 and DAPI (blue) for ddPCR. Arrow indicates the picked single cell.

**Supplementary Figure 4.**
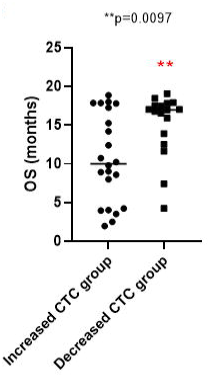
Kaplan–Meier plot for the changes of EpCAM-positive CTCs after chemotherapy. Kaplan–Meier survival curve was stratified by the comparative changes (increased or decreased) of EpCAM^+^ CTC counts after chemotherapy (*p=0.0152).

**Supplementary Figure 5.**
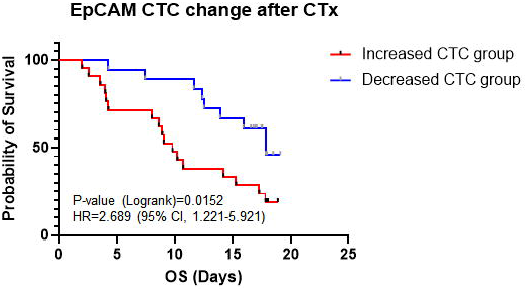
Overall survival depending on the changes of CTCs after chemotherapy. The relative change of CTCs between pre-CTx and post-CTx (increased or decreased) was evaluated with overall survival of PDAC patients (**p=0.0097).

## < Source Data Legends >

**Figure 1-source data 1. Uncropped blot with label for cytokeratin, EpCAM and Plectin-1 in PDAC cell lines**.

The expression of EpCAM, pan cytokeratin (CK), and Plectin-1 were determined by immunoblotting in PDAC cell lines (Capan-1, AsPC1, PK59, HPAFII, PANC1, MIA-PaCa2 cells) and WBCs. GAPDH was used as an internal control marker.

**Figure 1-source data 2**. Raw blot for cytokeratin (CK)

**Figure 1-source data 3**. Raw blot for EpCAM

**Figure 1-source data 4**. Raw blot for GAPDH

**Figure 1-source data 5**. Raw blot for plectin-1

**Table 1 &Figure 3-source data 1. All dataset for enrolled 52 patients**

A total of 52 patients with PDAC was enrolled, and their clinical information and CTC counts were collected. The first sheet includes data for a total of 52 patients, and the second sheet includes data for 39 patients with paired CTC counts after chemotherapy.

## REFERNECES

1. Ryan DP, Hong TS, Bardeesy N. Pancreatic adenocarcinoma. The New England journal of medicine. 2014;371(11):1039–49.

2. Kleeff J, Korc M, Apte M, La Vecchia C, Johnson CD, Biankin AV, et al. Pancreatic cancer. Nat Rev Dis Primers. 2016;2:16022.

3. Rahib L, Smith BD, Aizenberg R, Rosenzweig AB, Fleshman JM, Matrisian LM. Projecting cancer incidence and deaths to 2030: the unexpected burden of thyroid, liver, and pancreas cancers in the United States. Cancer research. 2014;74(11):2913–21.

4. Siegel RL, Miller KD, Jemal A. Cancer statistics, 2020. CA: a cancer journal for clinicians. 2020;70(1):7–30.

5. Oberstein PE, Olive KP. Pancreatic cancer: why is it so hard to treat? Therap Adv Gastroenterol. 2013;6(4):321–37.

6. Harsha HC, Kandasamy K, Ranganathan P, Rani S, Ramabadran S, Gollapudi S, et al. A compendium of potential biomarkers of pancreatic cancer. PLoS Med. 2009;6(4):e1000046.

7. Ferrone CR, Finkelstein DM, Thayer SP, Muzikansky A, Fernandez-delCastillo C, Warshaw AL. Perioperative CA19-9 levels can predict stage and survival in patients with resectable pancreatic adenocarcinoma. Journal of clinical oncology : official journal of the American Society of Clinical Oncology. 2006;24(18):2897–902.

8. Hidalgo M. Pancreatic cancer. The New England journal of medicine. 2010;362(17):1605–17.

9. Nagrath S, Jack RM, Sahai V, Simeone DM. Opportunities and Challenges for Pancreatic Circulating Tumor Cells. Gastroenterology. 2016;151(3):412–26.

10. Keller L, Pantel K. Unravelling tumour heterogeneity by single-cell profiling of circulating tumour cells. Nat Rev Cancer. 2019;19(10):553–67.

11. Fan JL, Yang YF, Yuan CH, Chen H, Wang FB. Circulating Tumor Cells for Predicting the Prognostic of Patients with Hepatocellular Carcinoma: A Meta Analysis. Cell Physiol Biochem. 2015;37(2):629–40.

12. Gasparri ML, Savone D, Besharat RA, Farooqi AA, Bellati F, Ruscito I, et al. Circulating tumor cells as trigger to hematogenous spreads and potential biomarkers to predict the prognosis in ovarian cancer. Tumour biology : the journal of the International Society for Oncodevelopmental Biology and Medicine. 2016;37(1):71–5.

13. Heitzer E, Haque IS, Roberts CES, Speicher MR. Current and future perspectives of liquid biopsies in genomics-driven oncology. Nat Rev Genet. 2019;20(2):71–88.

14. Cristofanilli M, Budd GT, Ellis MJ, Stopeck A, Matera J, Miller MC, et al. Circulating tumor cells, disease progression, and survival in metastatic breast cancer. The New England journal of medicine. 2004;351(8):781–91.

15. Naito T, Tanaka F, Ono A, Yoneda K, Takahashi T, Murakami H, et al. Prognostic impact of circulating tumor cells in patients with small cell lung cancer. J Thorac Oncol. 2012;7(3):512–9.

16. Olmos D, Arkenau HT, Ang JE, Ledaki I, Attard G, Carden CP, et al. Circulating tumour cell (CTC) counts as intermediate end points in castration-resistant prostate cancer (CRPC): a single-centre experience. Annals of oncology : official journal of the European Society for Medical Oncology / ESMO. 2009;20(1):27–33.

17. Ferreira MM, Ramani VC, Jeffrey SS. Circulating tumor cell technologies. Mol Oncol. 2016;10(3):374–94.

18. Ignatiadis M, Lee M, Jeffrey SS. Circulating Tumor Cells and Circulating Tumor DNA: Challenges and Opportunities on the Path to Clinical Utility. Clinical cancer research : an official journal of the American Association for Cancer Research. 2015;21(21):4786–800.

19. Joosse SA, Gorges TM, Pantel K. Biology, detection, and clinical implications of circulating tumor cells. EMBO Mol Med. 2015;7(1):1–11.

20. DiPardo BJ, Winograd P, Court CM, Tomlinson JS. Pancreatic cancer circulating tumor cells: applications for personalized oncology. Expert Rev Mol Diagn. 2018;18(9):809–20.

21. Bausch D, Thomas S, Mino-Kenudson M, Fernandez-del CC, Bauer TW, Williams M, et al. Plectin-1 as a novel biomarker for pancreatic cancer. Clinical cancer research : an official journal of the American Association for Cancer Research. 2011;17(2):302–9.

22. Song BG, Kwon W, Kim H, Lee EM, Han YM, Kim H, et al. Detection of Circulating Tumor Cells in Resectable Pancreatic Ductal Adenocarcinoma: A Prospective Evaluation as a Prognostic Marker. Frontiers in Oncology. 2021;10(3457).

23. Kang HM, Kim GH, Jeon HK, Kim DH, Jeon TY, Park DY, et al. Circulating tumor cells detected by lab-on-a-disc: Role in early diagnosis of gastric cancer. PloS one. 2017;12(6):e0180251.

24. Kim TH, Lim M, Park J, Oh JM, Kim H, Jeong H, et al. FAST: Size-Selective, Clog-Free Isolation of Rare Cancer Cells from Whole Blood at a Liquid-Liquid Interface. Anal Chem. 2017;89(2):1155–62.

25. Waters AM, Der CJ. KRAS: The Critical Driver and Therapeutic Target for Pancreatic Cancer. Cold Spring Harb Perspect Med. 2018;8(9).

26. Poruk KE, Gay DZ, Brown K, Mulvihill JD, Boucher KM, Scaife CL, et al. The clinical utility of CA 19-9 in pancreatic adenocarcinoma: diagnostic and prognostic updates. Curr Mol Med. 2013;13(3):340–51.

27. Yu M, Bardia A, Wittner BS, Stott SL, Smas ME, Ting DT, et al. Circulating breast tumor cells exhibit dynamic changes in epithelial and mesenchymal composition. Science (New York, NY). 2013;339(6119):580–4.

28. Poruk KE, Valero V, 3rd, Saunders T, Blackford AL, Griffin JF, Poling J, et al. Circulating Tumor Cell Phenotype Predicts Recurrence and Survival in Pancreatic Adenocarcinoma. Annals of surgery. 2016;264(6):1073–81.

29. Grover PK, Cummins AG, Price TJ, Roberts-Thomson IC, Hardingham JE. Circulating tumour cells: the evolving concept and the inadequacy of their enrichment by EpCAM-based methodology for basic and clinical cancer research. Annals of oncology : official journal of the European Society for Medical Oncology / ESMO. 2014;25(8):1506–16.

30. Rofi E, Vivaldi C, Del Re M, Arrigoni E, Crucitta S, Funel N, et al. The emerging role of liquid biopsy in diagnosis, prognosis and treatment monitoring of pancreatic cancer. Pharmacogenomics. 2019;20(1):49–68.

31. Gires O, Stoecklein NH. Dynamic EpCAM expression on circulating and disseminating tumor cells: causes and consequences. Cell Mol Life Sci. 2014;71(22):4393–402.

32. Kim MY, Oskarsson T, Acharyya S, Nguyen DX, Zhang XH, Norton L, et al. Tumor self-seeding by circulating cancer cells. Cell. 2009;139(7):1315–26.

33. Janni WJ, Rack B, Terstappen LW, Pierga JY, Taran FA, Fehm T, et al. Pooled Analysis of the Prognostic Relevance of Circulating Tumor Cells in Primary Breast Cancer. Clinical cancer research : an official journal of the American Association for Cancer Research. 2016;22(10):2583–93.

34. Cohen SJ, Punt CJ, Iannotti N, Saidman BH, Sabbath KD, Gabrail NY, et al. Relationship of circulating tumor cells to tumor response, progression-free survival, and overall survival in patients with metastatic colorectal cancer. Journal of clinical oncology : official journal of the American Society of Clinical Oncology. 2008;26(19):3213–21.

35. Hiltermann TJN, Pore MM, van den Berg A, Timens W, Boezen HM, Liesker JJW, et al. Circulating tumor cells in small-cell lung cancer: a predictive and prognostic factor. Annals of oncology : official journal of the European Society for Medical Oncology / ESMO. 2012;23(11):2937–42.

36. Sefrioui D, Blanchard F, Toure E, Basile P, Beaussire L, Dolfus C, et al. Diagnostic value of CA19.9, circulating tumour DNA and circulating tumour cells in patients with solid pancreatic tumours. British journal of cancer. 2017;117(7):1017–25.

37. Kulemann B, Rösch S, Seifert S, Timme S, Bronsert P, Seifert G, et al. Pancreatic cancer: Circulating Tumor Cells and Primary Tumors show Heterogeneous KRAS Mutations. Sci Rep. 2017;7(1):4510.

38. Kim JE, Lee KT, Lee JK, Paik SW, Rhee JC, Choi KW. Clinical usefulness of carbohydrate antigen 19-9 as a screening test for pancreatic cancer in an asymptomatic population. J Gastroenterol Hepatol. 2004;19(2):182–6.

39. McGuigan A, Kelly P, Turkington RC, Jones C, Coleman HG, McCain RS. Pancreatic cancer: A review of clinical diagnosis, epidemiology, treatment and outcomes. World journal of gastroenterology : WJG. 2018;24(43):4846–61.

40. Gerlinger M, Rowan AJ, Horswell S, Math M, Larkin J, Endesfelder D, et al. Intratumor heterogeneity and branched evolution revealed by multiregion sequencing. The New England journal of medicine. 2012;366(10):883–92.

41. Brungs D, Minaei E, Piper AK, Perry J, Splitt A, Carolan M, et al. Establishment of novel long-term cultures from EpCAM positive and negative circulating tumour cells from patients with metastatic gastroesophageal cancer. Sci Rep. 2020;10(1):539.

42. Kim H, Lim M, Kim JY, Shin SJ, Cho YK, Cho CH. Circulating Tumor Cells Enumerated by a Centrifugal Microfluidic Device as a Predictive Marker for Monitoring Ovarian Cancer Treatment: A Pilot Study. Diagnostics (Basel). 2020;10(4).

